# START Diabetes Prevention: A Multi-Level Strategy for Primary Care Clinics

**DOI:** 10.1101/2024.06.10.24308653

**Authors:** Eva Tseng, Jill A. Marsteller, Jeanne M. Clark, Nisa M. Maruthur

## Abstract

**Background:** Prediabetes, a high-risk state for developing diabetes, affects more than 1 in 3 adults nationally. However, <5% of people with prediabetes are receiving any treatment for prediabetes. Prior intervention studies for increasing prediabetes treatment uptake have largely focused on individual barriers with few multi-level interventions that address clinician- and system-level barriers.

**Objective:** To measure the effectiveness of a multi-level intervention on uptake of prediabetes treatment in a primary care clinic.

**Design:** Pragmatic study of the START (Screen, Test, Act, Refer and Treat) Diabetes Prevention intervention.

**Participants:** The START Diabetes Prevention intervention was implemented in a suburban primary care clinic outside of Baltimore compared to a control clinic in the same area over a 12-month period.

**Intervention:** START Diabetes Prevention intervention included a structured workflow, shared decision-making resources and electronic health record clinical decision support tools.

**Main Measures:** Uptake of prediabetes treatment, defined as Diabetes Prevention Program referral, metformin prescription and/or medical nutrition referral within 30 days of any PCC visit.

**Key Results:** We demonstrated greater uptake of preventive treatment among patients with prediabetes in the intervention clinic vs. control clinic receiving usual care (11.6% vs. 6.7%, p<0.001). More patients in the intervention vs. control clinic reported their PCC discussed prediabetes with them (60% vs. 48%, p=0.002) and more felt overall that they understood what their doctor was telling them about prediabetes and that their opinion was valued. The START Diabetes Prevention Strategy had greater acceptability and usefulness to PCCs at the study end compared to baseline.

**Conclusions:** A low-touch multi-level intervention is effective in increasing prediabetes treatment uptake. The intervention was also acceptable and feasible for clinicians, and enhanced patient understanding and discussions of prediabetes with their clinicians.

## Introduction

One in 3 U.S. adults have prediabetes (1), a condition that increases the risk of type 2 diabetes with a 5-year risk of up to 50% (2). Prediabetes also increases the risk of stroke, heart attack and microvascular complications like neuropathy and nephropathy even before the onset of type 2 diabetes (3–5). Fortunately, effective treatments that prevent or delay type 2 diabetes include intensive lifestyle programs like the Diabetes Prevention Program (DPP) and metformin. The DPP randomized controlled trial demonstrated a reduction in diabetes incidence by 58% at 3 years and 27% at 15 years in the intensive lifestyle intervention group and by 31% at 3 years and 18% at 15 years in the metformin group, compared to the placebo group (6, 7).

Despite these effective treatments, translating these findings into practice and increasing the reach of diabetes prevention strategies to people with prediabetes have been difficult. Current data suggest that <5% of eligible people with prediabetes participate in a DPP and <5% of patients with prediabetes take metformin (8, 9). Individual barriers to the uptake of prediabetes treatment include a lack of knowledge about effective treatments and difficulty maintaining the lifestyle changes need to prevent diabetes (10). Clinician barriers include low knowledge about DPPs and a lack of time to counsel patients about prediabetes (11–14). System-level barriers include insufficient access and availability of diabetes prevention resources such as DPPs, nutrition counseling, and weight loss programs (14).

The American Medical Association (AMA) and Centers for Disease Prevention and Control (CDC) have partnered to recommend a system-wide strategy called Prevent Diabetes STAT™ (15) that addresses the following domains in the prediabetes care process: identify and screen at-risk individuals early, engage in shared decision discussion about treatment, and support patients in self-management and ongoing follow-up to monitor for progression. To our knowledge, few system-wide interventions have been studied with most focused on improving the DPP referral process (16–18) while missing the other domains in the prediabetes care process.

Our objective is to conduct a pragmatic study of a multi-level intervention, called START Diabetes Prevention that is based on Prevent Diabetes STAT™, to increase prediabetes treatment uptake in a primary care clinic.

## Methods

### Setting and Participants

The START diabetes prevention intervention was implemented in a suburban, academically affiliated primary care clinic outside of Baltimore with 11,000 patient visits in 2023 compared to a control clinic in the same area with 8,500 patient visits in 2023. A total of 18 clinicians practice in the intervention clinic (total includes 3 physicians who left and 1 physician and 1 nurse practitioner who started during the intervention period). Clinical staff members included 4 medical office coordinators, 3 registered nurses, and 3 medical assistants. A total of 6 physicians practice in the control clinic (1 physician left during intervention period).

All patients with prediabetes were eligible to participate in the study. Patients with prediabetes were identified using the EHR which contains a “Prediabetes Registry” that automatically identifies individuals meeting the following criteria: diagnosis of prediabetes on the problem list, a lab value in the prediabetes range (hemoglobin A1c 5.7-6.4%, fasting glucose of 100-125mg/dL, 2-hour glucose from oral glucose tolerance test of 140-199mg/dL), or ICD diagnosis code for prediabetes (R73.01, R73.02, R73.03) with a look-back period of 365 days.

The Institutional Review Board of Johns Hopkins School of Medicine reviewed and gave ethical approval this study. Since the study intervention was deemed to be part of routine care, we were not required to obtain informed consent of clinicians and patients. We obtained consent prior to the patient and clinician surveys. This study is registered at clinical trials.gov (NCT05265312).

### Intervention Design and Development

We used the Translating Evidence into Practice (TRiP) framework (19), an established implementation science method for translating evidence into real world practice incorporating 4 stages: 1) summarize the evidence, 2) identify local barriers to implementation, 3) measure performance, and 4) ensure all patients receive the intervention by implementing the 6 E’s (engage, educate, execute, evaluate, embed, and expand). A full description of how we applied the TRiP Framework to the first 3 phases of intervention design and development are described in a separate paper (20).

The START (“Screen, Test, Act, Refer, and Treat”) Diabetes Prevention clinical pathway is described in more detail elsewhere (20) and briefly summarized below. The steps of this clinical pathway were outlined in a clinician treatment algorithm that was printed and distributed to all clinicians in the intervention clinic (Appendix 1).

1. Screen: identify patients at-risk for prediabetes/diabetes who are due for screening using an automated EHR clinical decision support tool (“Diabetes Screening Best Practice Advisory” alert) that flags eligible patients using criteria from the 2021 US Preventive Services Task Force guidelines (21). This alert pops up during visit encounters if the patient meets the eligibility criteria (age 35-70 years and BMI ≥25kg/m^2^).
2. Test: clinician can order one of several laboratory tests (fasting glucose, hemoglobin A1c, or point-of-care hemoglobin A1c) which appear in the alert described in the “Screen” step.
3. Act: if a patient is diagnosed with prediabetes based on glycemic test results, the clinician and patient may have a shared decision-making discussion about diabetes preventive treatment options using a Prediabetes Decision Aid, which was developed and pilot tested by O’Brien et al. (22). The one-page Prediabetes Decision Aid pamphlet, included in the clinician treatment algorithm (Appendix 1), displays visually using 3 icon arrays the absolute risk of developing type 2 diabetes when participating in an intensive lifestyle intervention like the DPP, taking metformin, or without any treatment. Each icon array has a picture of 100 people, and color shading is used to represent the number of people likely to develop type 2 diabetes under each treatment scenario over a 3-year timeframe based on results from the DPP randomized controlled trial (6).
4. Refer/Test: based on the above conversation, clinicians may decide to refer patients to evidence-based diabetes prevention interventions (DPP and/or medical nutrition therapy) and/or prescribe metformin. While not directly studied in the DPP trial, individualized medical nutrition therapy (MNT) has been demonstrated to improve glycemic control in people with prediabetes (23, 24). Each of these orders are included in the EHR orders set (“prediabetes orders smartset”) we developed.
5. Follow-up: regular follow-up between clinician and patients to evaluate treatment and progress. In the clinician treatment algorithm, we suggested a follow-up visit timeframe of every 3-6 months to continue discussions about prediabetes knowing there may be insufficient time for this topic to be addressed at every visit. We also recommended clinicians recheck laboratory tests at least every 12 months consistent with guidelines (25).

### Implementation of Intervention

Several weeks prior to the study start date, we provided an in-person and virtual training session open to all PCCs in the intervention clinic. We recorded this training session and shared the recording and training slides with PCCs who could not attend the session. The training session covered the evidence supporting the START Diabetes Prevention intervention, reviewed all the steps in the clinical pathway, and shared the resources available in the implementation toolkit (e.g., clinician treatment algorithm, patient handout). As part of this implementation toolkit, we encouraged clinicians to take an online educational module (offered 0.75 Continuing Education credit hours) designed to teach clinicians how to engage with patients in discussions about diabetes prevention using established motivational interviewing techniques (26). We did not notify PCCs in the control clinic about the intervention study nor did they receive any specific training.

During each week of the 12-month active intervention period (5/30/22-6/2/23), we ran a report and reviewed a list of patients scheduled to have a routine (non-urgent) visit with their PCC the following week in the intervention clinic. For this list of identified patients, we sent a message through MyChart, the patient portal messaging system, a message encouraging the patient to read the attached informational handout about prediabetes and to bring it to their appointment with their PCC to discuss further (Appendix 2). If the patient did not have an active MyChart account, we sent the message and handout to their listed email address. Patients in the control clinic did not receive this informational handout and received usual care. If patients responded that they did not have prediabetes, we reviewed their chart to confirm this finding and added them to a list of ineligible patients.

As described earlier, we encouraged PCCs in the intervention clinic to use the START Diabetes Prevention clinical pathway and toolkit components. However, we did not require them to use any of the printed resources nor any of the EHR clinical decision support tools.

### Evaluation of Program

#### Main Measures and Data Collection

The primary outcome of our study was placement of a referral to a DPP, referral to medical nutrition therapy, and/or metformin prescription within 30 days after a PCC visit (yes/no) during the intervention period. Our secondary outcomes include prediabetes ICD diagnosis code placed at PCC visit (yes/no), medical nutrition therapy visit completion among those with a referral (yes/no), follow-up PCC visit within 7 months of index PCC visit (yes/no), glycemic laboratory test order placed (yes/no) and test completion (yes/no), and weight loss from baseline weight at index PCC visit to final weight ever recorded during intervention period (achieve ≥5% body weight loss, yes/no). The 12-month baseline period was from 5/31/22-5/29/22 and the 12-month active intervention period was from 5/30/22-6/2/23.

Other sociodemographic data were obtained from EHR data, including age, sex, race/ethnicity, and insurance plan at their index visit V0, defined as the patient’s first visit with the PCC during the intervention period. Vitals and laboratory tests, including BMI, blood pressure, fasting glucose, and hemoglobin A1c, were also obtained from EHR within a 90-day window around V0.

#### PCC Surveys

We obtained feedback from PCCs at baseline, 6 and 12 months through an online survey using REDCap (Research Electronic Data Capture). Survey domains included expectancy, instrumentality and adoption of intervention components, valence, acceptability, and actionability of START Diabetes Prevention. In the 12 months survey, we asked additional questions related to how the intervention could be improved.

#### Patient Surveys

We ran weekly reports to determine which eligible patients with prediabetes completed a visit with their PCC in the prior week. Three to five days after each completed visit with their PCC, we sent patients in both the intervention and control clinics an email with a link to a survey in REDCap. To increase the survey response rate, we sent up to 3 email reminders and attempted to call patients at least once in the 2 weeks following the survey being sent. Survey domains asked about whether their PCC discussed prediabetes with them, perceptions of that discussion (felt understood, opinion valued, felt doctor’s concern), what that discussion entailed, their attitudes toward prediabetes, and lifestyle changes they may be making.

#### EHR Clinical Decision Support Tools

We assessed the adoption of two EHR CDS tools: 1) DPP Best Practice Advisory-alerts clinicians when eligible patients are due for screening, 2) prediabetes orders smartset-bundle of orders related to prediabetes management.

#### Analysis

For the analysis, we considered the patient’s first visit with the PCC during the intervention period to be the index visit (V0). We confirmed that patients in the analytic cohort had prediabetes based on the “Prediabetes Registry” criteria discussed earlier. We excluded patients who became deceased during the active intervention period. We required patients to have had at least 1 PCC visit in the baseline period and at least PCC visit in the active intervention period since the intervention occurs in the context of a visit. There were 2 PCC departures in the intervention clinic and 1 PCC departure in the control clinic. Patients remained in the analytic cohort if they were reassigned and transferred their care to another PCC in the same clinic. Otherwise, they were excluded from the analysis.

We conducted descriptive analyses comparing the sociodemographic characteristics of patients in each clinic at time V0. We compared means and proportions using t-tests and chi-squared tests, respectively. For the primary outcome of DPP referral, MNT referral and/or metformin prescription within 30 days of any PCC visit, we compared the proportion of patients with this outcome during the 12-month intervention period using t-test for comparison between clinics. For the other secondary outcomes, we evaluated the proportion of patients with this outcome during the 12-month intervention period using t-test for comparison between clinics.

For the PCC surveys, we were unable to compare paired data at baseline to 12 months due to small number of respondents and little variation in responses. For questions with a Likert scale on agreement, we dichotomized the answers by combining agree and strongly agree vs. neither agree nor disagree, disagree and strongly disagree. For questions with a Likert scale on frequency, we also dichotomized the answers. For the patient surveys, we examined surveys where patients reported their PCC had discussed prediabetes with them then compared the proportion of patient visits where each domain occurred, using t-test for comparison between clinics.

For the DPP Best Practice Advisory, we evaluated the percentage of PCC visits where the alert fired and the percentage of PCC visits where the alert fired and a DPP referral was placed.

For the prediabetes orders smartset, we evaluated the percentage of PCC visits where the orders smartset was used.

All data analysis was performed used SAS or Stata 15.1 (StataCorp LLC, College Station, Texas).

## Results

Over the 12-month intervention period, 873 patients were seen in the intervention clinic and 1037 patients were seen in the control clinic. In the intervention clinic, patients with prediabetes were slightly older (62.5 years vs. 60.7 years, p=0.006) but similar in proportion of female sex (62.4% vs. 60%, p=0.23) and race/ethnicity (Table 1). The intervention clinic had slightly more patients with Johns Hopkins Employee Health Plan and Medicare insurance whereas the control clinic had more patients with other insurance plans. Mean BMI, fasting plasma glucose, hemoglobin A1c and blood pressure were similar in both clinics.

**Table 1.** Baseline characteristics of patients at initial PCC visit, comparing intervention to control clinic.

| Characteristic | Intervention clinic<br>(n=873) | Control clinic<br>(n=1037) | p-value |
| --- | --- | --- | --- |
| Age, mean (SD) | 62.52 (13.6) | 60.74 (14.6) | 0.006 |
| Age category, n (%) |  |  | 0.02 |
| 18-26 years | 14 (1.6) | 14 (1.4) |  |
| 27-44 years | 81 (9.3) | 149 (14.4) |  |
| 45-64 years | 354 (40.6) | 411 (39.6) |  |
| 65-74 years | 260 (29.8) | 282 (27.2) |  |
| 75+ years | 164 (18.8) | 181 (17.5) |  |
| Female sex, n (%) | 545 (62.4) | 617 (59.5) | 0.23 |
| Race, n (%) |  |  | 0.20 |
| Asian | 62 (7.1) | 93 (9.0) |  |
| Black | 220 (25.2) | 280 (27.0) |  |
| White | 544 (62.3) | 604 (58.2) |  |
| Other | 33 (3.8) | 49 (4.7) |  |
| Unknown/Declined to answer | 14 (1.6) | 11 (1.1) |  |
| Hispanic ethnicity, n (%) | 19 (2.2) | 26 (2.5) | 0.89 |
| Insurance plan, n (%) |  |  | <0.001 |
| Commercial | 412 (47.2) | 487 (47.0) |  |
| Johns Hopkins Employee Health Plan | 106 (12.1) | 100 (9.6) |  |
| Medicare | 314 (36.0) | 326 (31.4) |  |
| Medicaid | 25 (2.86) | 17 (1.6) |  |
| Self-pay | 7 (0.8) | 10 (1.0) |  |
| Other | 9 (1.03) | 97 (9.4) |  |
| BMI, mean (SD) | 30.55 (6.8) | 30.82 (6.8) | 0.42 |
| BMI category, n (%) |  |  | 0.65 |
| <25 | 152 (19.8) | 177 (18.9) |  |
| 25.0 to <30 | 259 (33.8) | 298 (31.8) |  |
| 30.0 to <35 | 186 (24.3) | 252 (26.9) |  |
| 35.0 to <40 | 107 (14.0) | 124 (13.3) |  |
| ≥40.0 | 62 (8.1) | 85 (9.1) |  |
| Fasting plasma glucose, mean (SD) | 97.18 (12.7) | 97.3 (13.0) | 0.89 |
| Fasting plasma glucose <100 mg/dL, n (%) | 275 (71.1) | 295 (67.4) | 0.57 |
| Fasting plasma glucose 100-109 mg/dL, n (%) | 66 (17.1) | 77 (17.6) |  |
| Fasting plasma glucose 110-125 mg/dL, n (%) | 35 (9.0) | 49 (11.2) |  |
| Fasting plasma glucose >125 mg/dL, n (%) | 11 (2.8) | 17 (3.9) |  |
| HbA1c, mean (SD) | 5.83 (0.3) | 5.84 (0.3) | 0.33 |
| HbA1c, <5.7% n (%) | 114 (19.8) | 97 (16.6) | 0.57 |
| HbA1c, 5.7-5.9% n (%) | 285 (49.4) | 301 (51.5) |  |
| HbA1c, 6-6.4% n (%) | 172 (29.8) | 180 (30.8) |  |
| HbA1c, >6.4% n (%) | 6 (1.0) | 7 (1.2) |  |
| Systolic BP (mmHg), mean (SD) | 129.77 (16.3) | 129.24 (16.9) | 0.52 |
| Diastolic BP (mmHg), mean (SD) | 78.08 (8.9) | 78.5 (11.0) | 0.38 |

For the primary outcome, 11.6% of patients in the intervention clinic had a DPP referral, MNT referral and/or metformin prescription within 30 days of any PCC visit during the intervention compared to 6.7% of patients in the control clinic day (p<0.001). Secondary outcomes included ICD diagnosis code of prediabetes placed at PCC visit, which was significantly higher in the intervention vs. control clinic, both at baseline and during the intervention period (Table 2). MNT visits occurred infrequently among patients with a referral placed, with no difference between the two clinics. The percentage of patients having at least one follow-up PCC visit in the 7 months after V0 was slightly higher in the control clinic vs. intervention clinic (56% vs. 50%, p=0.01). Glycemic tests ordering and completion rates were higher in the intervention clinic, both during the baseline and intervention periods, but the between-group differences were not significant. The percentage of patients achieving 5% body weight loss from baseline weight at V0 was similar in both clinics (p=0.14).

**Table 2.**
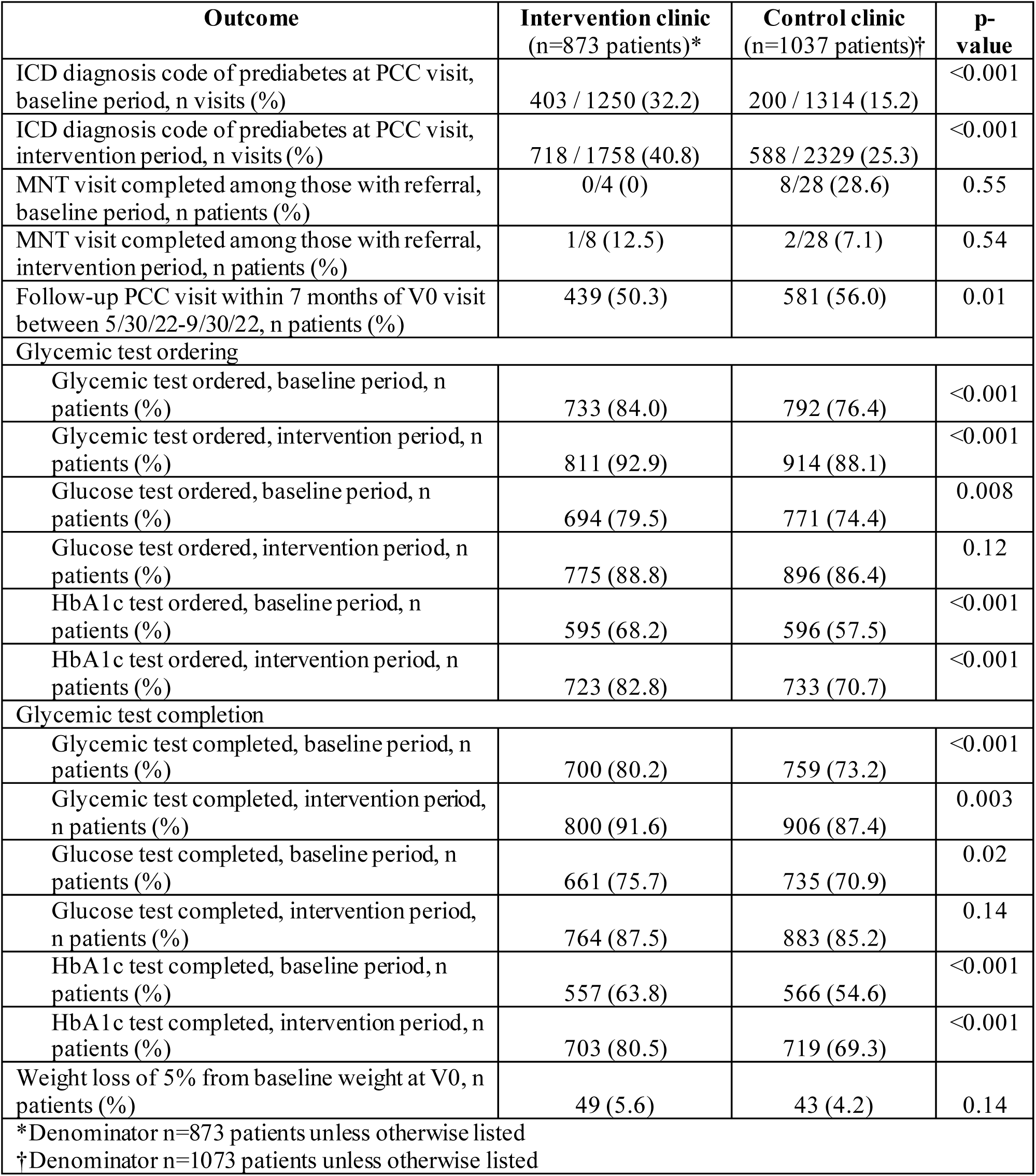
Secondary outcomes, including ICD diagnosis of prediabetes, medical nutrition therapy visits, follow-up PCC visits, and glycemic test ordering and completion, comparing the intervention and control clinics.

| **Outcome** | **Intervention clinic**<br>**(n=873 patients)\*** | **Control clinic**<br>**(n=1037 patients)†** | **p-value** |
| --- | --- | --- | --- |
| ICD diagnosis code of prediabetes at PCC visit, baseline period, n visits (%) | 403 / 1250 (32.2) | 200 / 1314 (15.2) | <0.001 |
| ICD diagnosis code of prediabetes at PCC visit, intervention period, n visits (%) | 718 / 1758 (40.8) | 588 / 2329 (25.3) | <0.001 |
| MNT visit completed among those with referral, baseline period, n patients (%) | 0/4 (0) | 8/28 (28.6) | 0.55 |
| MNT visit completed among those with referral, intervention period, n patients (%) | 1/8 (12.5) | 2/28 (7.1) | 0.54 |
| Follow-up PCC visit within 7 months of V0 visit between 5/30/22-9/30/22, n patients (%) | 439 (50.3) | 581 (56.0) | 0.01 |
| **Glycemic test ordering** |  |  |  |
| Glycemic test ordered, baseline period, n patients (%) | 733 (84.0) | 792 (76.4) | <0.001 |
| Glycemic test ordered, intervention period, n patients (%) | 811 (92.9) | 914 (88.1) | <0.001 |
| Glucose test ordered, baseline period, n patients (%) | 694 (79.5) | 771 (74.4) | 0.008 |
| Glucose test ordered, intervention period, n patients (%) | 775 (88.8) | 896 (86.4) | 0.12 |
| HbA1c test ordered, baseline period, n patients (%) | 595 (68.2) | 596 (57.5) | <0.001 |
| HbA1c test ordered, intervention period, n patients (%) | 723 (82.8) | 733 (70.7) | <0.001 |
| **Glycemic test completion** |  |  |  |
| Glycemic test completed, baseline period, n patients (%) | 700 (80.2) | 759 (73.2) | <0.001 |
| Glycemic test completed, intervention period, n patients (%) | 800 (91.6) | 906 (87.4) | 0.003 |
| Glucose test completed, baseline period, n patients (%) | 661 (75.7) | 735 (70.9) | 0.02 |
| Glucose test completed, intervention period, n patients (%) | 764 (87.5) | 883 (85.2) | 0.14 |
| HbA1c test completed, baseline period, n patients (%) | 557 (63.8) | 566 (54.6) | <0.001 |
| HbA1c test completed, intervention period, n patients (%) | 703 (80.5) | 719 (69.3) | <0.001 |
| Weight loss of 5% from baseline weight at V0, n patients (%) | 49 (5.6) | 43 (4.2) | 0.14 |
| *Denominator n=873 patients unless otherwise listed |  |  |  |
| †Denominator n=1073 patients unless otherwise listed |  |  |  |

### PCC Survey

The baseline survey was completed by 9 out of 11 PCCs in the intervention clinic whereas the 12-month survey was completed by 11 out of 16 PCCs (denominators are not the same due to turnover) (Table 4). Baseline survey respondents had a range of years in practice (25% ≤5 years, 25% 6-10 years, 25% 11-20 years, 25% 21+ years) and clinical effort (56% ≤25% Full-Time Equivalent, FTE; 11% 25.1-50% FTE; 33% 50.1-7% FTE).

In general, PCCs agreed that the treatment algorithm and Prediabetes Decision Aid were implementable and helped to improve quality of care, involve patients in their own care, and improve patient clinical outcomes (Table 4). Overall, the START Diabetes Prevention Strategy had greater acceptability and usefulness to PCCs at the study end compared to baseline. Regarding adoption of CDS tools, while use increased from baseline to 12 months, rates were still low for the different components (data not shown).

### Patient Survey

There were 319 surveys (n= 231 unique patients) completed in the intervention clinic and 254 surveys (n=196 unique patients) completed in the control clinic (Table 5). More patients in the intervention vs. control clinic reported that their doctor had discussed prediabetes with them at their recent visit (60% vs. 48%, p=0.002). Slightly more patients in the intervention vs. control clinic reported understanding of what their doctor told them about prediabetes, discussing options and ways to address prediabetes, and felt their doctor’s concern about prediabetes, although the difference between clinics was not statistically significant.

### CDS Tool Adoption in Intervention Clinic

The DPP Best Practice Advisory alert fired in 43% of PCC visits during the intervention period (Table 3). A DPP referral was placed in 4% of visits where the DPP Best Practice Advisory alert fired. The prediabetes orders smartset was used in only 1% of visits in both the baseline and intervention periods.

**Table 3.** Adoption of prediabetes clinical decision support tools in the intervention clinic.

| **CDS Tool** | **N visits (%)** |
| --- | --- |
| DPP Best Practice Advisory alert firing at PCC visit, intervention period | 760 / 1758 (43.2) |
| DPP referral placed during PCC visit, intervention period | 71 / 1758 (4.0) |
| DPP referral placed during PCC visit where DPP Best Practice Advisory alert fired, intervention period | 27 / 760 (3.6) |
| Prediabetes orders smartset used, baseline period | 14 / 1250 (1.1) |
| Prediabetes orders smartset used, intervention period | 21 / 1758 (1.2) |
| BPA, Best Practice Advisory; CDS, clinical decision support; DPP, Diabetes Prevention Program, PCC, primary care clinician |  |

**Table 4.** PCC survey responses at baseline and 12 months.

| **Agreement with Survey Domain** | **Baseline**<br>**(N=9)**<br>**N (%)** | **12-month**<br>**Survey**<br>**(N=11)**<br>**N (%)** |
| --- | --- | --- |
| **PCC treatment algorithm\*†** |  |  |
| Exerting effort will help to implement | 7 (88) | 11 (100) |
| Help improve quality of care for patients with prediabetes | 8 (100) | 10 (91) |
| Help involve patients in own care | 7 (88) | 11 (100) |
| Help improve patient clinical outcomes | 8 (100) | 10 (91) |
| **Prediabetes Decision Aid\*†** |  |  |
| Exerting effort will help to implement | 7 (88) | 11 (100) |
| Help improve quality of care for patients with prediabetes | 8 (100) | 10 (91) |
| Help involve patients in own care | 7 (88) | 10 (91) |
| Help improve patient clinical outcomes | 8 (100) | 9 (82) |
| **Acceptability‡** |  |  |
| START Diabetes Prevention Strategy works for my patients | 4 (44) | 9 (82) |
| START Diabetes Prevention Strategy adds to workload in unacceptable way | 0 | 0 |
| The more I incorporate START Diabetes Prevention Strategy, the better care I can provide patients with prediabetes | 5 (56) | 8 (73) |
| START Diabetes Prevention Strategy is not applicable for my patients with prediabetes | 0 | 0 |
| **Actionability/Usefulness‡** |  |  |
| I will implement changes in my practices based on START Diabetes Prevention Strategy | 6 (67) | 10 (91) |
| Amount of time required to understand START Diabetes Prevention Strategy is excessive | 1 (11.1) | 0 |
| START Diabetes Prevention Strategy is relevant to patient-related decisions I will make | 8 (89) | 9 (82) |
| I have the ability to change care to improve prediabetes management using START Diabetes Prevention Strategy | 6 (67) | 9 (82) |
| I understand the purpose of START Diabetes Prevention Strategy | 9 (100) | 11 (100) |
| START Diabetes Prevention Strategy doesn't apply to large proportion of my patients with prediabetes | 1 (11) | 0 |
| I have a clear understanding of how to use START Diabetes Prevention Strategy to discuss treatment options | 8 (89) | 9 (82) |
| START Diabetes Prevention Strategy is too complex | 1 (11) | 0 |
| *Dichotomized as agree (strongly agree, agree, slightly agree) vs. disagree (strongly disagree, disagree, slightly disagree, neither agree nor disagree) |  |  |
| †n=8 respondents to these questions |  |  |
| ‡Dichotomized as agree (strongly agree, agree) vs. disagree (strongly disagree, disagree, neither agree nor disagree) |  |  |

**Table 5.** Patient survey responses in the intervention vs. control clinic.

| Agreement with Survey Domain* | Intervention clinic<br>(N=319 visits)<br>N (%) | Control clinic<br>(N=254 visits)<br>N (%) | p-value |
| --- | --- | --- | --- |
| Discussion with PCC |  |  |  |
| Understood what doctor told me about prediabetes | 185/191 (97) | 111/120 (93) | 0.08 |
| Felt my opinion was valued when doctor talked to me about prediabetes | 179/191 (94) | 106/117 (91) | 0.31 |
| We discussed options and ways to address prediabetes | 170/190 (89) | 102/119 (86) | 0.32 |
| Felt my doctor's concern about prediabetes | 173/189 (92) | 108/120 (90) | 0.65 |
| Attitude towards prediabetes |  |  |  |
| I am worried about having prediabetes | 134/192 (70) | 92/121 (76) | 0.23 |
| I am motivated to take steps to reduce risk of getting diabetes | 181/191 (95) | 117/120 (98) | 0.24 |
| I feel confident in my ability to manage prediabetes | 156/190 (82) | 98/120 (82) | 0.92 |
| I feel able to meet challenge of controlling my prediabetes | 151/186 (81) | 97/119 (82) | 0.94 |
| *Dichotomized as agree (strongly agree, agree) vs. disagree (strongly disagree, disagree, neither agree nor disagree) |  |  |  |

## Discussion

In this pragmatic study of a multi-level intervention in primary care called START Diabetes Prevention, we demonstrated a small but significant increase in prediabetes treatment uptake in the intervention versus control clinic. Supported by our prior work, we applied the TRiP framework (19) to guide 3 phases of intervention design and development (20) to operationalize the Prevent Diabetes STAT™ strategy (15) for addressing the prediabetes care process. PCCs felt that the START Diabetes Prevention intervention was acceptable and feasible, improving their care of patients with prediabetes without adding to their workload. Furthermore, more patients in the intervention clinic versus control clinic had a better understanding of and felt that their opinion was valued in their discussions with their PCCs about prediabetes.

To our knowledge, few system-wide interventions for prediabetes have been studied with most focusing on improving the DPP referral process (16, 17) while missing the other domains in the prediabetes care process. One study conducted a cluster randomized trial in which intervention clinicians at a large academic family medicine clinic received a brief 1-hour training about prediabetes and the DPP and received a daily EHR report about patients eligible for diabetes screening and/or met prediabetes criteria. Clinicians in the intervention clusters referred 6.9% of patients with prediabetes to the DPP compared to 1.5% of patients in the control clusters (16). In a different study implementing clinical decision support tools to increase DPP referral at the Henry Ford Health System, the study team conducted a pre-post evaluation showing that among the 4,930 patients eligible for the DPP, 293 patients were referred during the 12-month intervention period compared to 20 referrals during the 6-month baseline period (17). Our results for uptake of DPP referrals were similar to both studies. However, our intervention was more comprehensive and incorporated and evaluated other domains in the prediabetes care process (e.g., follow-up PCP visits, glycemic testing and completion). We also gathered patient-reported outcomes, which the other two studies did not, adding to the current limited literature on system-wide interventions for prediabetes.

There are several strengths to our study including our intervention components were low touch and low-cost, although we did not conduct a formal cost analysis to demonstrate this due to the pilot nature of this study. Implementation costs included training time and delivery of shared-decision making discussions with patients, which occur routinely as part of primary care. We were able to incorporate clinical decision support tools to support screening, referral and follow-up of patients, but our data shows that adoption of these tools were low and this finding is not surprising and common in other similar types of interventions. Therefore, even without these clinical decision support tools, these interventions can be easily translated and adopted in other settings to increase prediabetes treatment uptake. Finally, we were able to prevent contamination as the intervention occurred in one clinic and was compared to a control clinic in a nearby location serving a similar patient population.

There are some limitations to our findings. Our intervention was implemented in a single primary care clinic, limiting the generalizability of our findings. There were other health system initiatives around diabetes prevention occurring around the time of this study that may have positively influenced treatment uptake, but these initiatives would affect both clinics equally. Finally, the intervention did not include other clinical staff members such as nurses or medical assistants, who can play in an important role in screening or follow-up of patients with prediabetes. Due to a staffing shortage in the intervention clinic, we were unable to incorporate other clinical staff members in the intervention, but recognize this arrangement can be an important way to augment the intervention.

In the future, we plan to implement this intervention across the health system in multiple primary care clinics and incorporate other clinical staff members in the intervention since we recognize that PCCs have many health issues to address in short visit. Other studies have utilized pharmacists (27) or medical assistants (22) to lead discussions about prediabetes treatment demonstrating a significant increase in uptake of DPP and/or metformin. This change will increase the likelihood of sustainability and dissemination.

In conclusion, our pragmatic study of a system-wide multi-level intervention, START Diabetes Prevention, was effective at increasing the uptake of evidence-based preventive treatment for prediabetes. The intervention was also acceptable and feasible for clinicians, and enhanced patients’ understanding and discussions of prediabetes with their clinicians.

## Funders

This study was funded by the National Institute of Diabetes and Digestive and Kidney Diseases [Dr. Eva Tseng, K23DK118205].

## Data Availability

All data produced in the present study are available upon reasonable request to the authors.

