## Appendices for "START Diabetes Prevention: A Multi-Level Strategy for Primary Care Clinics"

### Engaging patients in START Diabetes Prevention

#### How to engage patients about preventing diabetes

- Ask patient: What do you know about what you can do to prevent diabetes?
- Ask permission: Would it be okay for me to tell you more about diabetes prevention?
    - If yes, Educate: Use information in centerfold to have conversation
    - If no, Ask Permission to revisit at next appointment

#### Why it's important to treat prediabetes

- Prediabetes costs $500 annually per person in medical costs
- People with prediabetes have early evidence of retinopathy, neuropathy and chronic kidney disease
- Regression from prediabetes to normoglycemia is associated with lower prevalence of microvascular disease

#### Strategies for preventing diabetes

- Discuss preventing diabetes is possible
- Discuss making simple healthy changes:
    - Losing 5% of body weight
    - Eat fewer processed foods, trans fats, sugary drinks and alcohol
    - Get moving: gradually work up to being active at moderate intensity 30 min, 5 days a week
- Track progress: people who track their food, activity and weight are more likely to reach their goals
- Get support from family and friends

#### Talking to patients about DPPs

- Research has shown that the program helps people lose weight and prevent diabetes (see guide)
- DPPs focus on losing 5% of body weight and increasing exercise to 150 min/week
- People receive group support from peers and a trained lifestyle coach
- Johns Hopkins DPPs are free and offered virtually or in-person
- People receive tools for making lifestyle changes like recipes, measuring cups, and a scale

#### Talking to patients about Metformin

- Research has shown that Metformin can delay or prevent diabetes (see guide)
- Metformin may be particularly beneficial for people with BMI ≥35kg/$m^2$, age <60, FBG ≥110mg/dL, HbA1c≥6%, or prior gestational diabetes
- Metformin does not have to be a lifelong medication, especially if people make lifestyle changes. Guidelines do not provide a recommendation on duration of treatment.

Appendix 1. Clinician treatment algorithm (bi-fold)

JOHNS HOPKINS MEDICINE

### START Diabetes Prevention Strategy

#### SCREEN AT-RISK ADULTS (USPSTF 2021 Guidelines)

Age 35-70 years with BMI ≥25kg/$m^2$
(BMI ≥23kg/$m^2$ if Asian)

**PREDIABETES?**
FPG 100-125mg/dL
OR
HbA1c 5.7-6.4%

**Discuss evidence-based strategies to prevent type 2 diabetes using guide (back)**

- Diabetes Prevention Program (group, 12 months)*
- Metformin*
- Medical nutrition therapy (1:1)

**Not interested?**
Reassess interventions at next visit

**NORMAL?**
Rescreen in 3 years

**Refer to Hopkins Diabetes Prevention Program**
Eligible if prediabetes by lab + BMI≥ 25
After referral is placed, DPP coordinator will call to screen and enroll

**Metformin**
Aim for ≥1000mg total as tolerated
Consider if BMI≥35, FBS≥110, A1c≥6%, h/o gestational DM, or failed lifestyle intervention

**Refer to Medical Nutrition Therapy**
Known coverage: EHP, Priority Partners
For all others, will need to check with insurer**

#### FOLLOW-UP

- **3-6 months visits** follow-up visit to readdress or evaluate progress
- **9-12 months labs** check at least annually

Reassess interventions at follow-up visit

***Prediabetes orders smartset contains all these orders***

* Strongest evidence for diabetes prevention are Diabetes Prevention Program and Metformin. Both Johns Hopkins DPP and MNT visits are available in-person or virtually.

** After patient schedules, Johns Hopkins Nutrition Dept. conducts financial clearance to confirm insurance coverage.

Be specific with ICD diagnosis codes for MNT referral; add relevant diagnoses as some may be covered over others.

### Guide to Discussing DPP vs. Metformin

**Know your risk of developing diabetes**

#### OUT OF 100 PEOPLE LIKE YOU WHO JOIN A **DIABETES PREVENTION PROGRAM**...

14 of the 100 people with prediabetes **WILL** develop diabetes in 3 years

86 of the 100 people with prediabetes will **NOT** develop diabetes in 3 years

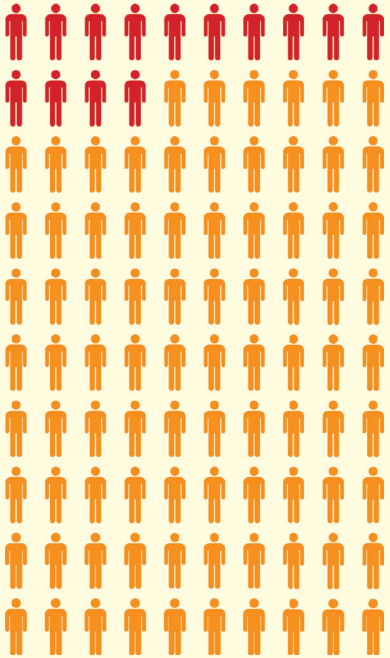

Joining a diabetes prevention program means:

- attending 1-hr group classes for a year
- losing 10 to 15 pounds (Johns Hopkins DPP participants lose over 5% body weight on average)
- eating healthy, being regularly active
- joining a program proven to work

#### OUT OF 100 PEOPLE LIKE YOU WHO TAKE **METFORMIN** TO PREVENT DIABETES...

22 of the 100 people with prediabetes **WILL** develop diabetes in 3 years

78 of the 100 people with prediabetes will **NOT** develop diabetes in 3 years

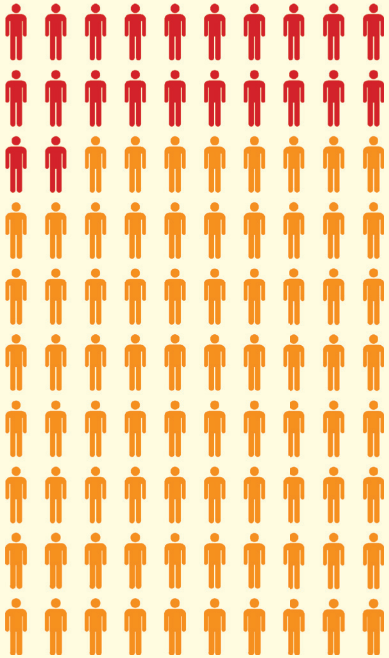

Taking Metformin to prevent diabetes means:

- taking a new medication once or twice a day until your doctor tells you to stop
- possibly losing 5 pounds
- possibly experiencing some side effects

#### OUT OF 100 PEOPLE LIKE YOU WHO MAKE **NO CHANGES**...

30 of the 100 people with prediabetes **WILL** develop diabetes in 3 years

70 of the 100 people with prediabetes will **NOT** develop diabetes in 3 years

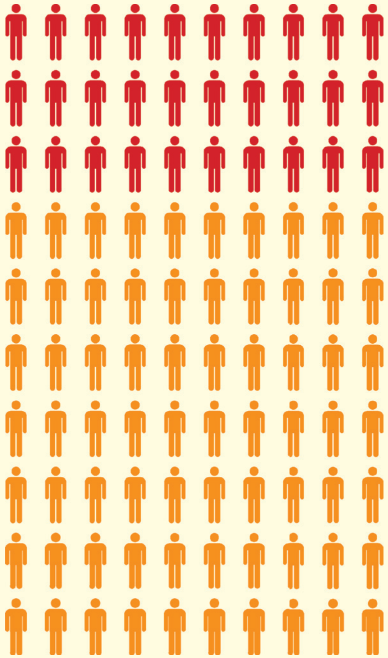

Developing diabetes means:

- pricking your finger regularly
- taking more than one medicine every day
- increasing your risk of blindness, dialysis, heart disease, stroke

*Adapted from Dr. Matthew O'Brien*

#### What is the Diabetes Prevention Program (DPP)?

- The DPP is a yearlong group lifestyle change program, developed by the Center for Disease Control and Prevention, delivered in person, online or virtually by trained lifestyle coaches.
- Involves 16 weekly sessions during the first six months. This is followed by one session a month for 6 months.
- You are eligible if you are 18 years and older, overweight, diagnosed with prediabetes or gestational diabetes, and no history of diabetes.
- You will learn how to increase physical activity, eat healthier, manage stress and overcome challenges to change.
- Covered by Medicare, Medicaid, and some private insurance companies. Programs at Johns Hopkins are free!
- Participants of the Johns Hopkins DPP have been successful. On average, participants lost over 5% of their body weight in 12 months!
- If interested in joining a Johns Hopkins DPP, let your primary care clinician know or contact the program by phone (410-614-2701) or by.

#### What do I need to know about metformin?

- Metformin is a medicine that is used to prevent and treat type 2 diabetes.
- Lowers your blood sugar levels by causing your liver to produce less glucose. It also prevents glucose from being absorbed in the belly.
- Some side effects of metformin include diarrhea, nausea, vomiting, and abdominal discomfort.
- Some people lose about 5 pounds when taking this medicine.
- It is important to continue making lifestyle changes if you start metformin.
- Talk to your primary care clinician if you have questions or are interested in starting metformin.

#### How can I learn more about prediabetes?

**For more information about prediabetes, go to the CDC website:**
*https://www.cdc.gov/diabetes/prevent-type-2/index.html*

**For more information about the Johns Hopkins Diabetes Prevention Program:**
*https://www.hopkinsmedicine.org/dpep/index.html*

**For more information on recipes and healthy food choices for prediabetes, go to the American Diabetes Association website:**
*https://www.diabetes.org/healthy-living/recipes-nutrition*

Appendix 2. Patient handout about prediabetes (bi-fold)

JOHNS HOPKINS
MEDICINE

### START Preventing Type 2 Diabetes

##### I talked to my doctor about prioritizing these steps to decrease my risk of diabetes:

- ❑ Joining a Diabetes Prevention Program
- ❑ Starting metformin at _______ dose
- ❑ Losing weight, starting with _______ pounds over ______ months
- ❑ Making healthier food choices
- ❑ Moving more, starting with _______ minutes per ___________
- ❑ Seeing a nutritionist
- ❑ Repeating lab tests in ____ months
- ❑ Having a visit within _____ months

**We want your feedback so we can help you control prediabetes better.**

**Look out for a survey that will be sent to your email!**

*5 minutes of your time for a $10 gift card!*

#### What is type 2 diabetes?

- Type 2 diabetes is a condition where the body does not respond normally to insulin (called insulin resistance). Eventually your pancreas cannot keep up. Your blood sugar then goes up leading to prediabetes and sometimes type 2 diabetes.
- People with type 2 diabetes may need to start medicine(s). They may need to watch out for serious health problems like heart disease, vision loss and kidney disease.

#### What can I do to prevent type 2 diabetes?

- Join a Diabetes Prevention Program (DPP). This is a yearlong lifestyle change program **(see back)**.
- Make healthier food choices. Eat fewer processed foods (white bread, crackers, pasta, sugary cereals). Decrease portion sizes.
- Increase physical activity. Set a goal of getting up to 30 minutes, 5 days/week of moderate exercise (like biking, jogging or brisk walking).
- Lose a little weight. As little as 5% of body weight can make a difference. For example, this is 10 pounds if you weigh 200 pounds or 12.5 pounds if you weigh 250 pounds.
- Talk with a nutritionist about the best food choices for you.
- Start metformin. This medicine has been shown to decrease your risk of type 2 diabetes **(see back)**.

#### What is prediabetes?

- Prediabetes is a condition where blood glucose levels are higher than normal but not high enough to be called type 2 diabetes.
- Prediabetes increases your risk of developing type 2 diabetes, heart attack, stroke and even dying earlier.

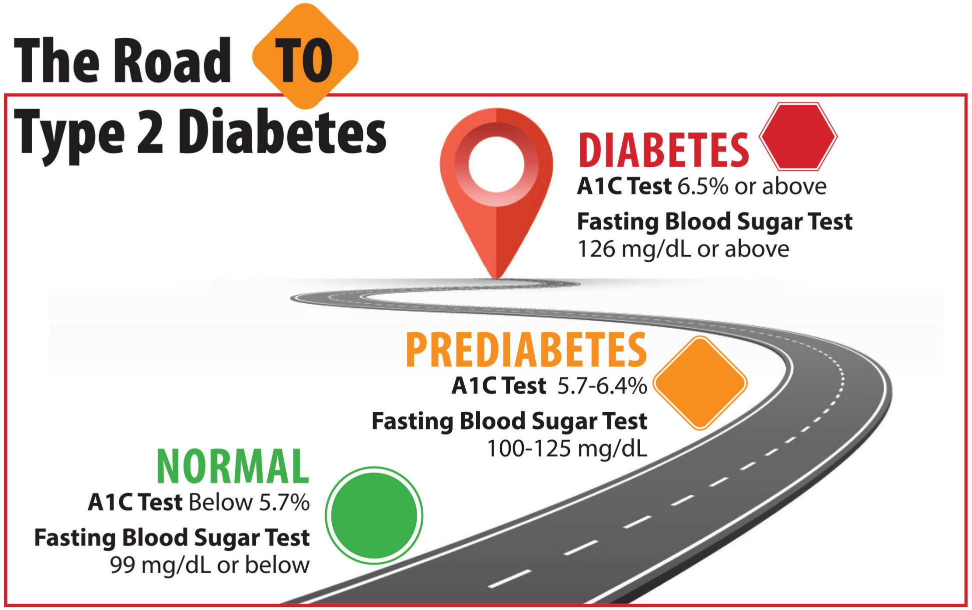


#### Know your risk of developing diabetes

OUT OF 100 PEOPLE LIKE YOU WHO JOIN A **DIABETES PREVENTION PROGRAM**...

14 of the 100 people with prediabetes **WILL** develop diabetes in 3 years

86 of the 100 people with prediabetes will **NOT** develop diabetes in 3 years

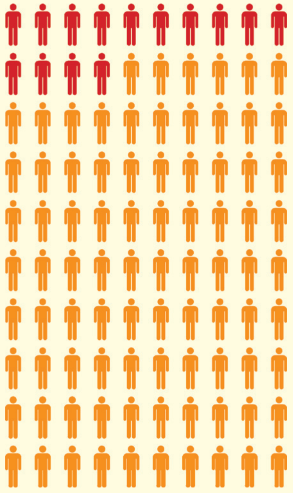

OUT OF 100 PEOPLE LIKE YOU WHO TAKE **METFORMIN** TO PREVENT DIABETES...

22 of the 100 people with prediabetes **WILL** develop diabetes in 3 years

78 of the 100 people with prediabetes will **NOT** develop diabetes in 3 years

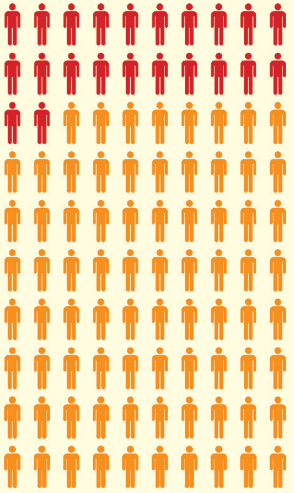

OUT OF 100 PEOPLE LIKE YOU WHO MAKE **NO CHANGES**...

30 of the 100 people with prediabetes **WILL** develop diabetes in 3 years

70 of the 100 people with prediabetes will **NOT** develop diabetes in 3 years

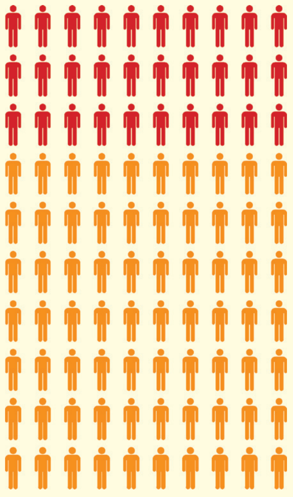

*Adapted from Dr. Matthew O'Brien*
